# Safety and efficacy of two immunization schedules with an inactivated SARS-CoV-2 vaccine in adults. A randomized non-inferiority clinical trial

**DOI:** 10.1101/2022.02.07.22270215

**Authors:** Katia Abarca, Carolina Iturriaga, Marcela Urzúa, Nicole Le Corre, Augusto Pineda, Carolina Fernández, Angélica Domínguez, Pablo A González, Susan M Bueno, Paulina Donato, Pilar Espinoza, Daniela Fuentes, Marcela González, Paula Guzmán, Paula Muñoz, Carlos M Pérez, Marcela Potin, Álvaro Rojas, José V González-Aramundiz, Nicolás MS Gálvez, Francisca Aguirre-Boza, Sofía Aljaro, Luis Federico Bátiz, Yessica Campisto, Marcela Cepeda, Aarón Cortes, Sofía López, María Loreto Pérez, Andrea Schilling, CoronaVac03CL Study Group, Alexis M Kalergis

## Abstract

**Background:** Several vaccines have been developed to control the COVID-19 pandemic. CoronaVac^®^ (Sinovac Life Sciences), an inactivated SARS-CoV-2 vaccine, has demonstrated safety and immunogenicity in previous studies, preventing severe COVID-19 cases. We further investigated the safety and efficacy of two immunization schedules of CoronaVac^®^ in a non-inferiority trial in healthy adults.

**Methods:** This is a multi-center and randomized clinical trial. Healthy adults were enrolled at eight centers in Chile. Participants were randomly assigned to two vaccination schedules, receiving two doses with either 14 (0-14) or 28 (0-28) days between each. 2302 participants were vaccinated. The primary safety and efficacy endpoints were solicited adverse events (AE) within 7 days after each dose and compared the number of cases of SARS-CoV-2 infection 14 days after the second dose between schedules, respectively.

**Findings:** The most frequent local AE was pain at the injection site, which was less frequent in participants aged ≥60 years. Other local AEs were reported in less than 5% of participants. The most frequent systemic AEs were headache, fatigue, and myalgia. The remaining AEs were minor allergic reactions and fever. Most AEs were mild and transient. There were no significant differences for local and systemic AE between schedules. No anaphylactic reactions or vaccine-related severe AEs were observed. 58 COVID-19 cases were confirmed, and all but two of them were mild. No differences were observed in protection between schedules.

**Interpretation:** CoronaVac^®^ is safe, especially in ≥60 years-old participants. Both schedules protected against COVID-19 hospitalizations.

**Funding:** MINSAL, Chile, CPC & IMII, Chile.

**Research in context:** *Evidence before this study:* CoronaVac^®^ (an inactivated SARS-CoV-2 vaccine) was approved on June 1^st^, 2021, by the WHO for its use in humans. Sinovac Life Sciences generated this vaccine in China and conducted phase 1/2 trials. Good safety, efficacy, and immunogenicity profiles were reported. The results from this study led to the use of CoronaVac^®^ in other countries, such as Brazil, Turkey, and Chile, with phase 3 trials being held on them.

*Added-value of this study:* This work compares the safety and efficacy of two immunization schedules with CoronaVac^®^, with each dose administrated two or four weeks after the first dose on healthy Chilean adults. To date, no studies showing the safety and efficacy of these two immunization schedules with CoronaVac^®^ in healthy adults in a population other than the Chinese have been published. We show that CoronaVac^®^ is safe and prevents hospitalization due to COVID-19 in both immunization schedules. No differences were found in the incidence of adverse events between both schedules, and no related severe adverse events were reported. These results give further insight into the immune response induced by CoronaVac^®^ and are relevant when deciding on the immunization schedule chosen for vaccination.

*Implications of all the available evidence:* The data reported here show that using either immunization schedule with two doses of CoronaVac^®^ protects against SARS-CoV-2. The data also indicate that CoronaVac^®^ does not induce severe adverse events in either immunization schedule, and the adverse events registered are mild and transient, confirming the safety of this vaccine.

## Background

In March 2020, the COVID-19 pandemic, a disease caused by the severe acute respiratory syndrome coronavirus 2 (SARS-CoV-2), was declared ^1^. Two years into this pandemic, more than 250 million cases have been diagnosed worldwide, and more than 5 million deaths can be related to COVID-19 ^2^. In Chile, since March 2020, 1.7 million laboratory-confirmed cases have been reported, and more than 38,000 deaths can be related to COVID-19 by December 2021 ^3^.

Initial COVID-19 outbreaks exhibited high morbidity and mortality in individuals over 60 years of age or with comorbidities, such as obesity, chronic pulmonary disease, cardiac disease, and immunosuppressed population ^4,5^. Antiviral drugs or immunomodulators have not been a successful treatment ^6^. Prophylactic strategies with drugs, such as hydroxychloroquine or ivermectin did not show any significant reduction in the risk of SARS-CoV-2 infection ^7^. Other treatments, such as post-exposure type I interferon prophylaxis, are still being evaluated ^8^.

Vaccination is an essential prophylactic strategy to prevent pathogen spreading and the disease caused by a viral infection ^9^. Early during the pandemic, the development of vaccines against SARS-CoV-2 was vigorously pursued. Different vaccine platforms were generated to prevent COVID-19, such as mRNA vaccines or viral vector-based vaccines ^10^. Among these, CoronaVac^®^ is an inactivated vaccine against SARS-CoV-2 developed in Vero cells (Sinovac Life Sciences, Beijing, China). Preclinical studies performed in mice, rats, and non-human primates demonstrated that this vaccine was immunogenic and induced anti-SARS-CoV-2 neutralizing antibodies ^11^. Moreover, partial or complete protection against pneumonia after a viral challenge was shown in primates ^11^. All these results led to human clinical trials. A phase I/II sequential clinical trial was performed, including 144 and 600 healthy adults aged 18 to 59 years, respectively ^12^. Two doses (3 and 6 µg) and two vaccination schedules (two doses separated by either two or four weeks) were evaluated. Results demonstrated that this inactivated vaccine was well tolerated with mild local adverse events after two doses ^12^. Although anti-SARS-CoV-2 neutralizing antibodies geometric mean titers (GMT) were lower when compared to convalescent patients, the vaccine induced a significant humoral response with both doses and schedules. A phase1/2 sequential clinical trial performed in healthy adults aged 60 years and older showed that CoronaVac^®^ was safe and well-tolerated in this particular population ^13^. Moreover, the 3 µg dose in the elderly group induced anti-SARS-CoV-2 neutralizing antibodies titers similar to those observed in adults aged 18-59 years. All these findings led to the emergency use of CoronaVac^®^ in China and supported the development of a phase 3 study to evaluate the efficacy of this inactivated vaccine ^14^.

Due to the availability of CoronaVac^®^ to the general public in Chile since January 2021, we adapted the initial placebo-controlled phase 3 clinical trial in adults to a non-inferiority clinical trial of two different immunization schedules, with the second dose administered either two (0-14) or four (0-28) weeks after the first one, with a planned 12 months of follow-up. This report includes the safety and efficacy of non-inferiority results acquired up to six months after the first dose.

## Methods

### Study design and participants

This trial (clinicaltrials.gov NCT04651790) is a multi-center, randomized clinical trial to evaluate two vaccination schedules of CoronaVac^®^, the Sinovac inactivated SARS-CoV-2 vaccine, in adults in Chile that included health care workers and community participants recruited at eight sites (six in the Metropolitan Region of Santiago and two in the Valparaiso Region). The study was approved by the sponsoring institution Ethical Committee (Comité Ético Científico Ciencias de la Salud UC, Pontificia Universidad Católica de Chile, ID 200708006), and each Institutional Ethical Committee of the other sites (Comité Ético Científico Universidad de Los Andes, Comité Ético Científico Facultad de Medicina Clínica Alemana, Universidad del Desarrollo, Comité Ético Científico Hospital Clínico Félix Bulnes, Comité Ético Científico Servicio de Salud Valparaíso-San Antonio, Comité Ético Científico Servicio de Salud Metropolitano Sur Oriente, Chile), and the Public Health Institute of Chile (ISP Chile, number N° 24204/20). This study was also conducted according to the current Tripartite Guidelines for Good Clinical Practices, the Declaration of Helsinki ^15^, and local regulations. An independent data and safety monitoring board reviewed the blinded safety and efficacy data.

Written informed consent was obtained from each participant before enrollment. After inclusion and exclusion criteria were met (a complete list of inclusion and exclusion criteria has been published previously ^16^), participants were randomly assigned to one of 2 open-label vaccination schedules, with either 14 (0-14) or 28 (0-28) days interval between doses, in a 1:1 ratio.

### Aims

The primary safety endpoint was to evaluate the frequency of AE occurring on the first 7 days after each dose of the vaccine in each vaccination schedule. The secondary endpoint was to determine the occurrence of SAE and events of special interest in both vaccination schedules during all the study.

The primary non-inferiority efficacy endpoint was to evaluate and compare the protection against confirmed SARS-CoV-2 infection of two vaccination schedules, starting two weeks after the second dose. Non-inferiority of 0-14 over 0-28 schedule was defined as a difference in the protection rate within a threshold of 15%. Secondary efficacy endpoints were to compare both vaccination schedules regarding hospitalized cases and deaths within the same period.

### Procedures

Demographic information, comorbidities, concomitant medications, and nutritional status were registered at enrolment and registered in a paper case report form (CRF) and an electronic CRF (eCRF). Blood samples and nasopharyngeal swabs were obtained for all participants prior to immunization to evaluate past or current SARS-CoV-2 infection. A urine test was performed on all female participants to assess potential pregnancies, an exclusion criterion. Participants were inoculated with 3 µg (600SU) of Coronavac^®^ and then kept in observation for 60 min after each dose to evaluate possible adverse events (AE). Immediate AE was defined as reporting the AE within this period. Then, participants, or their representative, if applicable, were instructed to register through a remote application any local and systemic solicited AE for 7 days after each dose and any other AE and concomitant medications until 28 days after the second dose. Non-immediate AEs were defined as those occurring after the first 60 minutes after vaccination. Serious adverse events (SAE), events of special interest (using the priority List of Events of Special Interest in COVID-19 vaccines by Brighton Collaboration) ^17^, relevant medications (immunosuppressive drugs, transfusions, and other vaccines), and symptoms of SARS-CoV-2 infection were collected throughout the entire study. The system sent daily reminders to all participants until day 28 after the second dose and then weekly until the end of the study. The severity of solicited AE was graded through a numeric scale of 1 to 4, based on the “Toxicity Grading Scale for Healthy Adult and Adolescent Volunteers Enrolled in Preventive Vaccine Clinical Trials” guide of the Food and Drug Administration of the United States (FDA) ^18^. The severity of the unsolicited clinical AE was classified through a numeric scale of 1 to 5, based on the “Common Terminology Criteria for Adverse Events - Version 5.0” guide by the United States National Cancer Institute (NCI /NIH) ^19^. The investigators determined a possible causal association between AE and vaccination according to a classification adapted from the “Uppsala Monitoring Center” of the World Health Organization ^20^. Personnel of the sites reviewed this information for accuracy and completeness and filled an AE or SAE form in the eCRF.

To determine the protection against confirmed SARS-CoV-2 infection for each vaccination schedule, participants were followed during the study to identify and register any SARS-CoV-2 infection (COVID-19). The definition of case surveillance for COVID-19 was stated by the WHO ^21^. Participants were instructed to register in the remote application and notify the sites through the mail, message, or phone call when they presented at least one of the symptoms for two days (suspicious case definition met). In these cases, a SARS-CoV-2 RT-qPCR was performed. A second sample was collected in the case of a negative RT-qPCR with persisting symptoms, and then a new RT-qPCR was performed. The investigators closely monitored participants who met the confirmed COVID-19 case definition (at least one symptom and a positive RT-qPCR), recording symptoms, severity, start and end dates, therapies, complications, hospitalizations, and admission to the ICU, use of mechanical ventilation, and outcome. The severity of the COVID19 symptoms was classified in 1 to 4 grades based on the guidelines “Toxicity Grading Scale for Healthy Adult and Adolescent Volunteers Enrolled in Preventive Vaccine Clinical Trials” from the United States Food and Drug Administration (FDA) and the “Common Terminology Criteria for Adverse Events - Version 5.0” guide by the United States National Cancer Institute (NCI /NIH) ^18,19^. The intensity of the condition was registered using the scale of clinical progression (Score 0 to 10), based on the WHO guidelines ^20^.

### Statistical analyses

Baseline characteristics of patients were compared by schedule: categorical variables were expressed as counts and percentages while numerical variables with mean and standard deviation (SD). Categorical variables were analyzed with Chi-square test or Fisher’s exact test; differences in means were tested using Student’s t-test; significance level was set at a more rigorous level of 0.01. The percentage of subjects that presented each solicited AE within the first 7 days was obtained for each schedule. The length of the event was presented as median and quantiles 10 and 90. Incidence of immediate and non-immediate AE was registered. The number of simultaneous non-immediate AEs were expressed as the sum of different AE and are shown as frequency and percentage by dose and schedule. Differences in the incidence of each AE by age were evaluated using Chi-square or Fisher’s exact test. The COVID-19 incidence, including only cases occurring 14 days after the second dose, was determined for each schedule and subgroups defined by sociodemographic or clinical characteristics. COVID-19-free survival was estimated using Kaplan-Meier analysis, and schedule curve differences were assessed using the Log Rank test. Cox’s regression was used to obtain age and gender-adjusted incidence rate ratios and their 95% confidence interval (CI). The proportional hazards assumption was met. For safety and efficacy, we looked at the non-inferiority of the 0-14 schedule over the 0-28 schedule, with a margin of 15%. Consequently, one-sided statistical tests were used where the rejection of the null hypothesis indicates non-inferiority of the 0-14 schedule over the 0-28 schedule. All statistical analyses were performed using SPSS 17.0.

## Results

### Demographics and participants

During November 29^th^, 2020, and April 9^th^, 2021, a total of 2,302 participants were vaccinated with the first dose of CoronaVac^®^. Of these participants, 1,090 were allocated to the 0-14 schedule and 1,212 to the 0-28 schedule. Safety and efficacy data derived from participants up to October 2021 are reported here, with a median (min-max) of follow-up of 6.5 (0.5-6.7) months for the 0-14 schedule and 6.9 (1.1-7.1) months for the 0-28 schedule. Safety information for 7 days after the first and second doses was available for 2,302 and 2,212 participants, respectively. These data were included in the safety analysis. Moreover, 2,205 participants had clinical information 14 days after the second dose and were included in the efficacy analysis (Figure 1).

**Figure 1.**
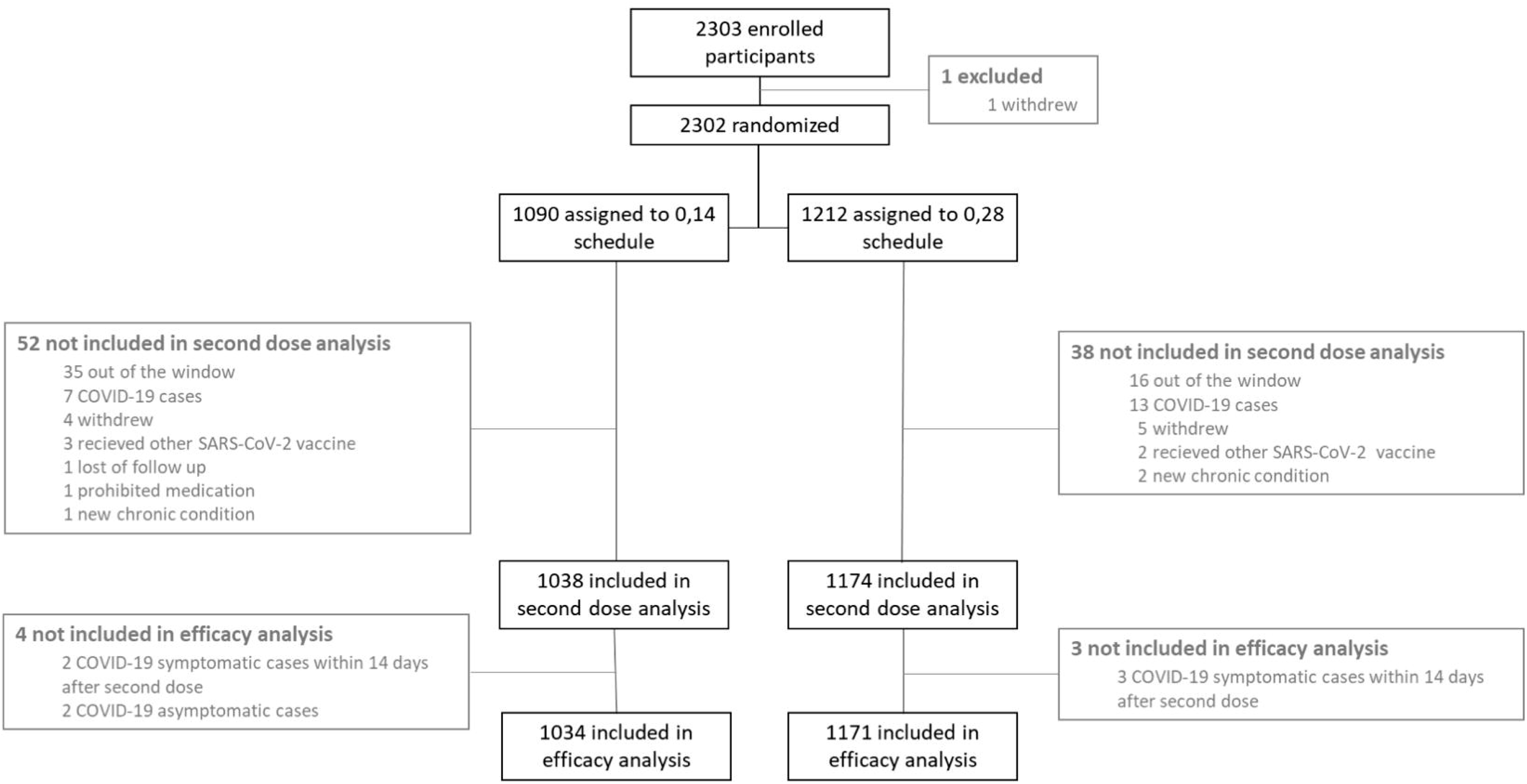
Study design for this Phase 3 trial with two different immunization schedules as of October 2021. This study aims to characterize the safety and efficacy elicited by two immunization schedules with CoronaVac^®^, with each dose separated by either 14 or 28 days.

Demographic characteristics and comorbidities of the population are shown in Table 1. No significant differences were observed between participants who received any of the two vaccination schedules, except a higher proportion of health workers in the 0-14 schedule; and a higher BMI and frequency of participants aged over 60 in the 0-28 schedule.

**Table 1.** Clinic and demographic characteristics of the study population at baseline.

|  | Total | Schedule 0-14 | Schedule 0-28 | p value (a) |
| --- | --- | --- | --- | --- |
|  | (n=2302) | (n=1090) | (n=1212) |  |
| Age; n (%) |  |  |  | **0.001** |
| 18-59 | 1616<br>(70.2) | 800 (73.4) | 816 (67.3) |  |
| 60-98 | 686 (29.8) | 290 (26.6) | 396 (32.7) |  |
| Sex; n (%) |  |  |  | 0.015 |
| Female | 1212<br>(52.6) | 603 (55.3) | 609 (50.2) |  |
| BMI; mean ± SD | 26.9 ± 4.5 | 26.6 ± 4.4 | 27.1 ± 4.6 | **0.003** |
| Ethnicity; n (%) |  |  |  | 0.128 |
| Hispanic or Latino | 2294<br>(99.7) | 1083 (99.4) | 1211 (99.9) |  |
| Chilean native | 4 (0.2) | 3 (0.3) | 1 (0.1) |  |
| Asian | 3 (0.1) | 3 (0.3) | 0 (0.0) |  |
| Black | 1 (0.0) | 1 (0.1) | 0 (0.0) |  |
| Health workers; n (%) |  |  |  | **<0.001** |
| Yes | 759 (33.0) | 459 (42.1) | 300 (24.8) |  |
| Comorbidities; n (%) |  |  |  |  |
| $\geq 1$ | 1042<br>(45.3) | 487 (44.7) | 555 (45.8) | 0.580 |
| Comorbidities; n (%) |  |  |  |  |
| Cardiovascular disease | 34 (1.5) | 14 (1.3) | 20 (1.7) | 0.468 |
| Asthma and COPD* | 148 (6.4) | 83 (7.6) | 65 (5.4) | 0.028 |
| Diabetes | 117 (5.1) | 44 (4.0) | 73 (6.0) | 0.030 |
| Insulin resistance | 180 (7.8) | 85 (7.8) | 95 (7.8) | 0.967 |
| Hypothyroidism | 248 (10.8) | 135 (12.4) | 113 (9.3) | 0.018 |
| Arterial hypertension | 415 (18.0) | 174 (16.0) | 241 (19.9) | 0.015 |
| Allergic rhinitis | 300 (13.0) | 137 (12.6) | 163 (13.4) | 0.531 |
| Thyroid disease | 251 (10.9) | 136 (12.5) | 115 (9.5) | 0.022 |
| Obesity | 478 (20.8) | 205 (18.8) | 273 (22.5) | 0.028 |
| Dyslipidaemia | 43 (1.9) | 18 (1.7) | 25 (2.1) | 0.467 |

### Safety parameters

During the first 60 minutes after vaccination, 1-2% of participants reported local pain at the administration site. The other AEs were recorded in even lower frequency (Supplementary Table 1). No anaphylactic reactions were observed. After this immediate period, a total of 882 local and 1,919 systemic solicited AEs were reported upon administration of the first dose. These AEs were reported in 32.1% and 41.5% of the vaccinated participants for the 0-14 and 0-28 schedules. A total of 867 local and 1395 systemic solicited AE were reported after the second dose. These AEs were reported in 31.2% and 32.9% of the vaccinated participants for the 0-14 and 0-28 schedules, respectively (Figure 1 and supplementary table 2). The 0-14 schedule showed no inferiority to the 0-28 schedule (p<0.0001) in the frequency of AEs (Supplementary Table 2).

After the first dose, 67.9% of the participants did not report any local AE, 26.6% reported only pain at the inoculation site, 0.9% reported pain and induration, 0.8% reported pain and local pruritus, and 0.6% reported only pruritus. All other combinations were found in less than 0.5% of the participants. After the second dose, 68.8% of the participants did not report any AE, 24.7% reported only pain at the inoculation site, 1.4% reported pain and induration, 0.8% reported pain, induration, and pruritus. All other combinations were found in less than 0.6% of the participants. Majority of the participants who presented any local AE after each dose reported one or two AEs (Supplementary Table 3).

After each dose, the most frequent solicited systemic AEs were headache, fatigue, and myalgia, reported in 20-26%, 12-17%, and 11-14% of the participants, respectively. The remaining systemic AEs were reported in less than 10% of the vaccinated participants. Notably, minor allergic reactions and fever were reported by less than 2% and 1% of the vaccinated participants, respectively (Figure 2 and Supplementary Table 2). The number of simultaneous systemic AEs is shown in Supplementary Table 3. The majority of the participants who presented any systemic AE reported one or two simultaneous AE. Most local and systemic AEs were mild, with 0.5% or fewer participants reporting grade 3 AEs after the first dose and 0.6% or less after the second dose. There were no reports of grade 4 AEs (Figure 2). The most frequent local and systemic AEs resolved in a median of 2 days (Supplementary Table 4). When comparing by age group, older participants (≥60 years old) showed less incidence of AEs than did younger participants (18-59 years old) (Supplementary Table 5).

**Figure 2.**
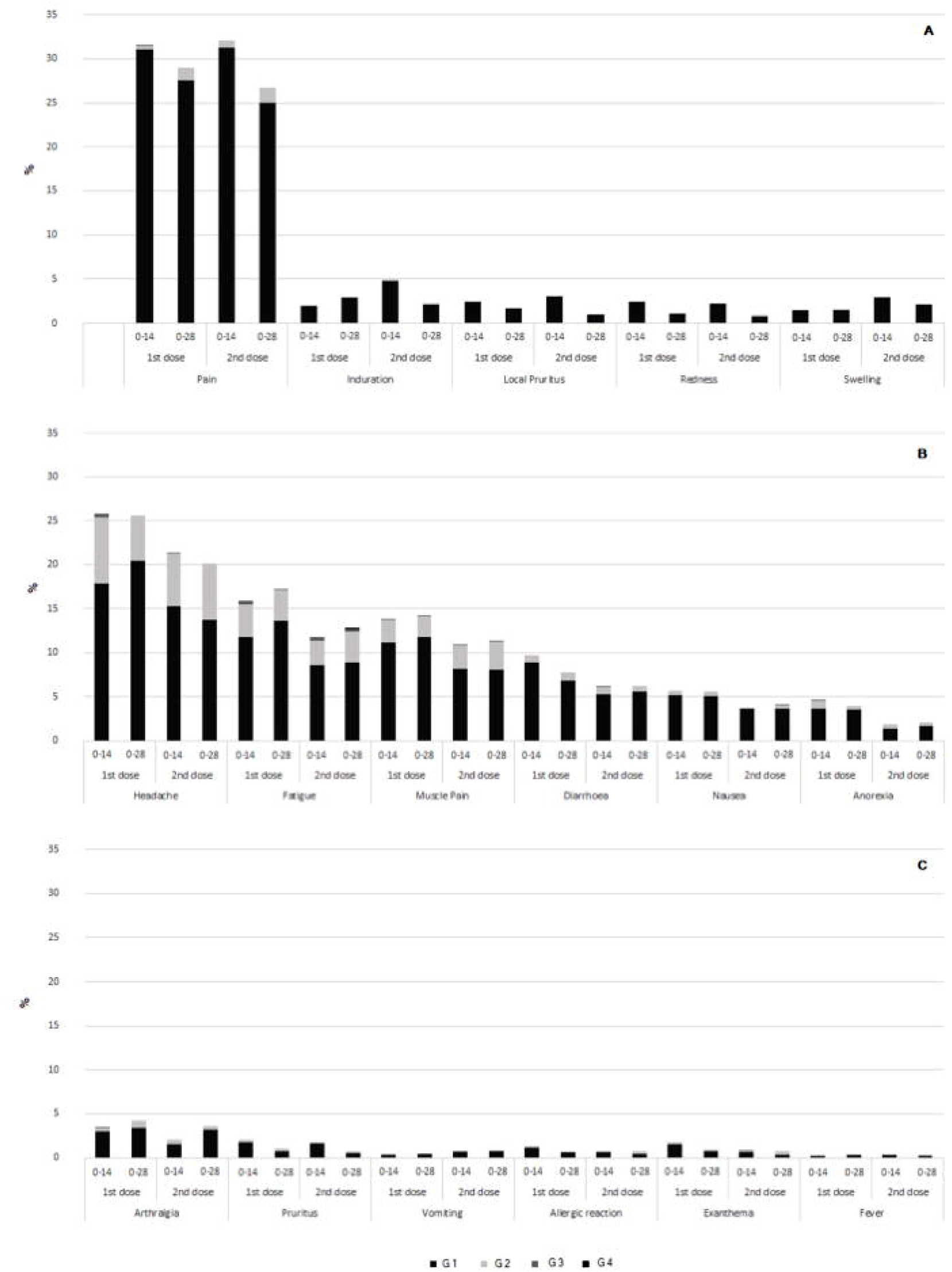
Frequency and severity of local and systemic adverse events by schedule and dose. Frequencies and severity grades are shown in percentage. (A) Local adverse events. (B & C) Systemic adverse events.

No vaccine-related SAE occurred, and 60 non-vaccine-related SAEs were reported, including 3 deaths: two sudden deaths, one due to acute myocardial infarction, one month after the second dose (male, between 50 and 60 years old), and other in a patient with a history of hepatic cirrhosis due to alcoholic liver disease, three months after the second dose (male between 60 and 70 years-old). The third death was due to gastric cancer Stage IV diagnosed five months after enrolment (female, between 70 and 80 years old). Five pregnancies have been reported in participants of the study, two of them during the first four weeks after the second dose, one twin and one single pregnancy. All had a negative pregnancy test and contraceptive use before each vaccine dose, and these participants are being followed-up by the investigators, with no obstetric nor perinatal complications reported to date. To date, one of the pregnancies concluded with the birth of healthy twins. No other events of special interest have occurred in the study.

### Vaccine efficacy

Upon 14 days after the administration of the second dose of CoronaVac^®^, 58 symptomatic and confirmed COVID-19 cases have been registered. The demographic and clinical characteristics of these COVID-19 cases are shown in Table 2. The vast majority of these cases were mild (Score 2) (94.8%), and just two participants were hospitalized. The first one was a male, aged over 60 years, BMI 28.0 (over-weighted), with arterial hypertension and bicuspid aorta. This participant exhibited COVID-19 symptoms 32 days after the second dose of a 0-28 schedule and was a confirmed close contact with a COVID-19 case. The participant developed atrial fibrillation and heart failure and required mechanical ventilation (Score 7) for six days and hospitalization for 20 days. The second participant is a male, aged over 60 years, BMI of 29.3 (over-weighted), in treatment for hypothyroidism. The second participant exhibited COVID-19 symptoms 122 days after the second dose of a 0-28 schedule, and no close contact with other COVID-19 cases. The participant received oxygen by nasal cannula (Score 5) for four days and was released after seven days of hospitalization. Both participants exhibited cough, dyspnea, and fatigue for more than seven weeks but ultimately recovered.

**Table 2.**
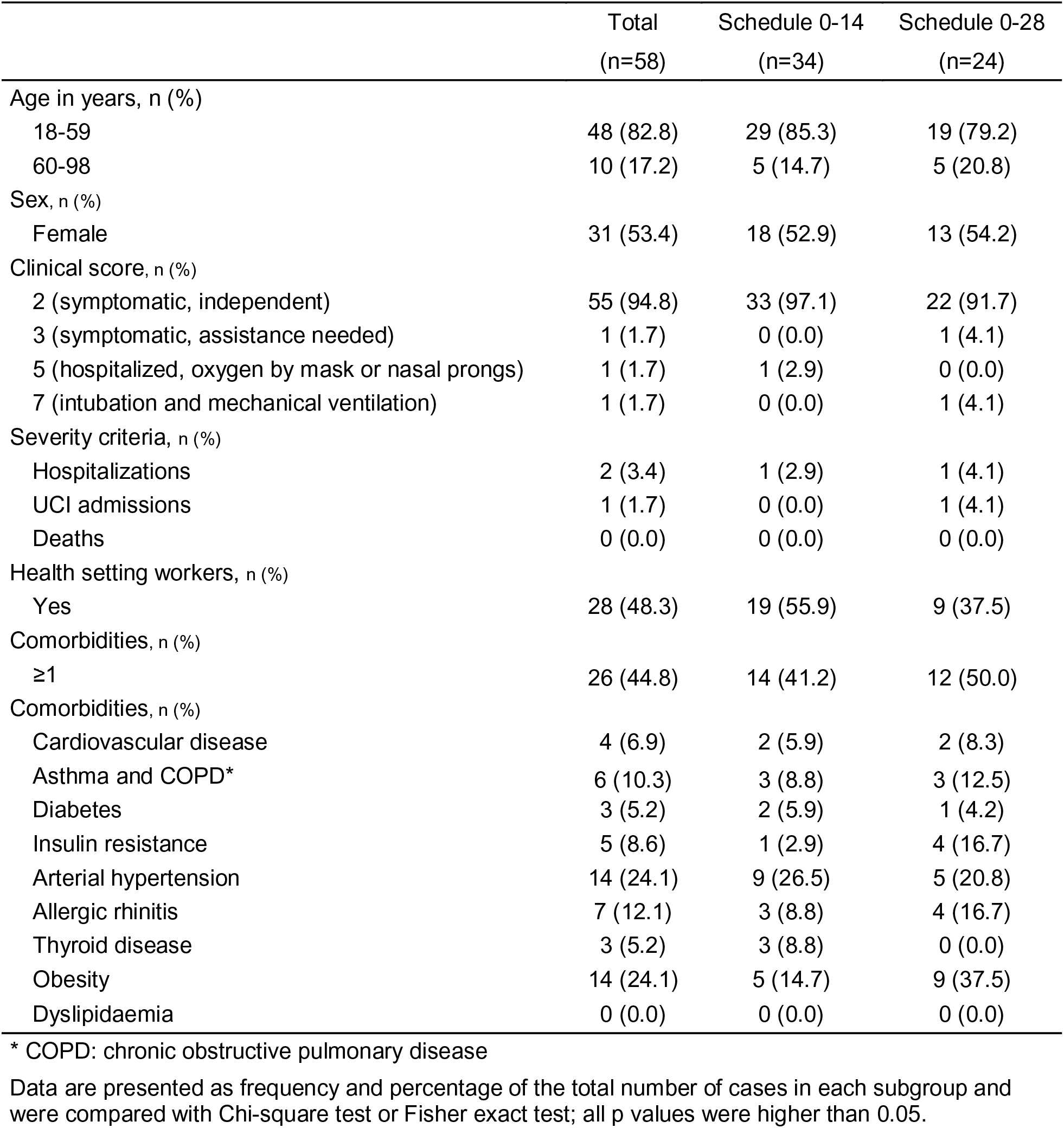
Characteristics of COVID-19 positive participants by immunization schedule.

|  | Total<br>(n=58) | Schedule 0-14<br>(n=34) | Schedule 0-28<br>(n=24) |
| --- | --- | --- | --- |
| Age in years, n (%) |  |  |  |
| 18-59 | 48 (82.8) | 29 (85.3) | 19 (79.2) |
| 60-98 | 10 (17.2) | 5 (14.7) | 5 (20.8) |
| Sex, n (%) |  |  |  |
| Female | 31 (53.4) | 18 (52.9) | 13 (54.2) |
| Clinical score, n (%) |  |  |  |
| 2 (symptomatic, independent) | 55 (94.8) | 33 (97.1) | 22 (91.7) |
| 3 (symptomatic, assistance needed) | 1 (1.7) | 0 (0.0) | 1 (4.1) |
| 5 (hospitalized, oxygen by mask or nasal prongs) | 1 (1.7) | 1 (2.9) | 0 (0.0) |
| 7 (intubation and mechanical ventilation) | 1 (1.7) | 0 (0.0) | 1 (4.1) |
| Severity criteria, n (%) |  |  |  |
| Hospitalizations | 2 (3.4) | 1 (2.9) | 1 (4.1) |
| UCI admissions | 1 (1.7) | 0 (0.0) | 1 (4.1) |
| Deaths | 0 (0.0) | 0 (0.0) | 0 (0.0) |
| Health setting workers, n (%) |  |  |  |
| Yes | 28 (48.3) | 19 (55.9) | 9 (37.5) |
| Comorbidities, n (%) |  |  |  |
| ≥1 | 26 (44.8) | 14 (41.2) | 12 (50.0) |
| Comorbidities, n (%) |  |  |  |
| Cardiovascular disease | 4 (6.9) | 2 (5.9) | 2 (8.3) |
| Asthma and COPD* | 6 (10.3) | 3 (8.8) | 3 (12.5) |
| Diabetes | 3 (5.2) | 2 (5.9) | 1 (4.2) |
| Insulin resistance | 5 (8.6) | 1 (2.9) | 4 (16.7) |
| Arterial hypertension | 14 (24.1) | 9 (26.5) | 5 (20.8) |
| Allergic rhinitis | 7 (12.1) | 3 (8.8) | 4 (16.7) |
| Thyroid disease | 3 (5.2) | 3 (8.8) | 0 (0.0) |
| Obesity | 14 (24.1) | 5 (14.7) | 9 (37.5) |
| Dyslipidaemia | 0 (0.0) | 0 (0.0) | 0 (0.0) |

\* COPD: chronic obstructive pulmonary disease
Data are presented as frequency and percentage of the total number of cases in each subgroup and were compared with Chi-square test or Fisher exact test; all p values were higher than 0.05.

A total of 34 and 24 cases of COVID-19 were registered in the 0-14 and the 0-28 schedule, respectively (p=0.083) (Table 2). Both schedules showed a high probability of being COVID-19-free: 96.7% (0-14) and 97.9% (0-28) (non-inferiority p-value <0.001). A Kaplan-Meier analysis showed that the probability of 0.98 for being COVID-19-free is achieved at day 91 for the 0-14 schedule and at day 133 for the 0-28 schedule. Although the COVID-19 incidence rate showed a slightly higher curve for the 0-14 than the 0-28 schedule, this difference was not statistically significant (log-rank test, p-value = 0.071) (Figure 3). The 0-14 schedule showed non inferiority to the 0-28 schedule when comparing COVID-19 incidence in different subpopulations defined by demographic and clinical characteristics (Supplementary Table 6).

**Figure 3.**
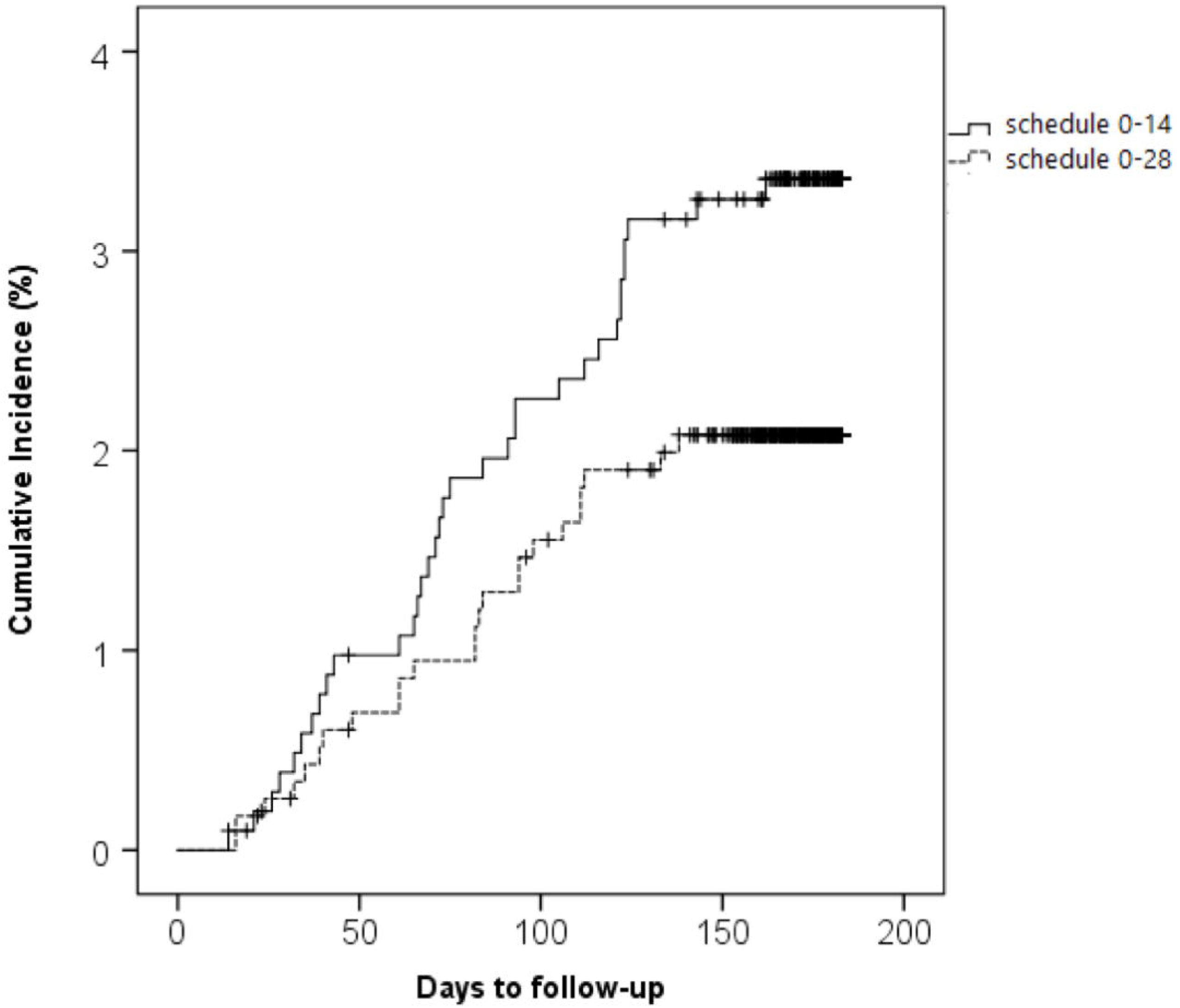
Cumulative incidence of COVID-19 infection by immunization schedule. Cumulative incidence for COVID-19 infections 14 days after administration of the second dose in two vaccination schedules (0-14 [solid line] and 0-28 [dashed line]). The X-axis shows days elapsed from the second dose to the event or censoring time. Censoring was set at the date of the third vaccination, retirement from the study, or reaching six months after the second dose, whichever occurred first.

The incidence of COVID-19 cases tends to be higher in health care workers compared with the general population for both immunization schedules (for 0-14 schedule, cases presented in 4.5 v/s 2.4% and for 0-28 in 3.1 v/s 1.7%); but these differences were not statistically significant. Also, the infection rate tends to be lower in ≥60 years old participants, but the significance level set for this analysis was not achieved (p=0.024). No statistical differences were observed in the frequency of COVID-19 cases between sex and comorbidities.

## Discussion

This non-inferiority trial demonstrated that the virus inactivated, CoronaVac^®^, given in two doses, with a 14 or 28 days interval between each dose, was safe, well-tolerated, and protective. A six-month surveillance showed a non-inferiority of the 0-14 over the 0-28 schedule in solicited AEs and confirmed COVID-19 cases. These results further support the safety and protective capacity for massive use of the Coronavac^®^ vaccine in adults, including participants older than 60 years old.

Regarding the safety of CoronaVac^®^, no vaccine-associated SAEs nor events of special interest were reported up until six months of follow-up in this cohort of over 2,300 adults. During phase III trials with adenovirus and mRNA-based vaccine formulations, four and one SAEs were associated with each vaccine, respectively ^22,23^. However, post-approval reports showed an increased incidence of vaccine-induced thrombotic thrombocytopenia, particularly for the adenovirus-based vaccine prototypes ^23^. Moreover, cases of vaccine-related myocarditis were observed in adolescents and young adults vaccinated with mRNA-based vaccine ^24^.

CoronaVac^®^ showed a low reactogenicity profile, with around 30% of vaccinated participants reporting local pain, less than 1% fever, and no significant allergic reactions. In this line, AEs reported on the phase III trials with CoronaVac^®^ performed in Turkey and Brazil were primarily mild and self-limited ^25,26^. The low reactogenicity profile of CoronaVac^®^ contrasts with the relatively high incidence of local and systemic AEs reported post-vaccination for other vaccine platforms, such as mRNA and adenoviral vectors, with pain observed in 50-80%, fever in 16-51%, fatigue up to 70%, and myalgias up to 60% of the participants ^22,27^.

Regarding the vulnerable population, 30% of the enrolled participants in this study were ≥60 years old, and 45% had chronic conditions. Lower frequencies of post-vaccination AEs were observed for the older age cohort compared to the younger participants. Consistently with this notion, only one elderly subject developed a fever after vaccination, a condition that could escalate in older people. Concordantly, in a nationwide cross-sectional study for side effects of CoronaVac^®^ performed in Turkey, younger age was a risk factor associated with an discrete increase of vaccine side effects ^26^. These features contribute to the confidence for the massive administration of this vaccine, especially in the most vulnerable populations.

Although the study design does not allow calculation of true efficacy for the vaccine due to the absence of a placebo arm, we demonstrate that in a scenario of high viral circulation ^3^, the vast majority of COVID-19 cases developed by vaccinated participants were only mild. Only two participants aged over 60 years required hospitalizations, and no deaths due to COVID-19 occurred as part of the study ^28^. These data are consistent with the immunogenicity results reported in Chile so far, showing that CoronaVac^®^ induces the secretion of specific IgG against the S1-RBD with neutralizing capacity, as well as the activation of T cells specific for SARS-CoV-2 antigens ^16^. Moreover, the wide use of this vaccine in the Chilean population was monitored by de Ministry of Health and has shown an effectiveness of 67,7% to prevent symptomatic COVID-19 cases and more than 85% to prevent severe COVID-19 cases and deaths due to SARS-CoV-2 infection ^29^.

Comparing the protective efficacy of two different vaccination schedules (0-14 v/s 0-28) against SARS-CoV-2 infection could help health authorities make evidence-based decisions for massive immunization against COVID-19. A more rapid schedule could lead to faster vaccination of the population, which could be relevant during an epidemic. It is essential to evaluate differences regarding immunogenicity, efficacy, and effectiveness between an accelerated schedule versus a standard four-week interval. Two previous reports with this vaccine showed a more robust immune response for the 0-28 schedule than for the 0-14 schedule ^12^. A phase I/II trial held in China showed higher neutralizing antibodies seroconversion rates for the 0-28 schedule compared to the 0-14 schedule ^12^.

Regarding efficacy, although we observed a trend toward higher efficacy for the 0-28 schedule compared to the 0-14 schedule, these differences were not statistically significant. A previous study that evaluated the efficacy of CoronaVac^®^ in a 0-14 schedule demonstrated that this parameter was higher in participants who received the two doses with an interval of over 21 days ^25^. An explanation for this apparent discrepancy is that in our study, the group included in the 0-14 schedule consisted mainly of healthcare workers, which are usually more exposed to the virus and therefore have higher risks of infection. Further studies with a more homogeneous population could contribute to addressing these questions.

Older age is a described risk factor related to COVID-19 severity ^30^, also observed in our study. Here, the two severe cases reported occurred just in older participants. However, the frequency of cases tended to be lower in this age group. This could be related to the most strict protective measures taken in this population and their lower mobility during the time of the study.

After a six-month follow-up, two doses of CoronaVac^®^ demonstrated to be well-tolerated, safe, and protective, particularly in a high-risk population. Regarding vaccination schedules, our data suggest that both a 0-14 and a 0-28 schedule show equivalent safety and efficacy results for this vaccine.

## Data Availability

All analyzed and raw data (masked to protect the information of volunteers) is available upon reasonable request to the corresponding authors through email after the publication of this article. A signed data access agreement will be requested to share the data. The study protocol is also available online and annexed to this article.

## Author contributions

**Conceptualization**: KA, PAG, SMB, JVG, AMK,

**Methodology**: KA, CI, MU, AP, CF, PD, PE, DF, MG, PG, PM, CMP, MP, AR.

**Investigation**: KA, CI, MU, AP, CF, PD, PE, DF, MG, PG, PM, CMP, MP, AR.

**Funding acquisition**: SMB, AMK.

**Project administration**: KA, PAG, SMB, AMK.

**Supervision**: KA, PAG, SMB, AMK.

**Writing – original draft**: KA, CI, MU, NLC, AD.

**Writing – review & editing**: KA, SMB, PAG, NMSG, AMK.

## Acknowledgments

We would like to thank the Ministry of Health, Government of Chile; Ministry of Science, Technology, Knowledge, and Innovation, Government of Chile; and The Ministry of Foreign Affairs, Government of Chile and the Chilean Public Health Institute (ISP). We also thank the Vice Presidency of Research (VRI), the Direction of Technology Transfer and Development (DTD), the Legal Affairs Department (DAJ) of the Pontificia Universidad Católica de Chile. We are also grateful to the Administrative Directions of the School of Biological Sciences and the School of Medicine of the Pontificia Universidad Católica de Chile for their administrative support. Special thanks to the independent data safety monitoring committee (members in the Supplementary Appendix (SA)) for their oversight and to the participants enrolled in the study for their participation and commitment to this trial. Members of the CoronaVac03CL Study Team are listed in the SA.

## References

1 Cucinotta D, Vanelli M. WHO Declares COVID-19 a Pandemic. Acta Biomed 2020; 91: 157–60.

2 WHO. Weekly Epidemiological Update on COVID-19. World Heal Organ 2020; : 1;4.

3 Ministerio de Salud G de C. Informe Epidemiológico N°164 Enfermedad por SARS-CoV-2. 2021. https://www.minsal.cl/wp-content/uploads/2021/12/Informe-Epidemiológico-164.pdf (accessed Jan 31, 2022).

4 Booth A, Reed AB, Ponzo S, et al. Population risk factors for severe disease and mortality in COVID-19: A global systematic review and meta-analysis. PLoS One 2021; 16: e0247461.

5 Zhou F, Yu T, Du R, et al. Clinical course and risk factors for mortality of adult inpatients with COVID-19 in Wuhan, China: a retrospective cohort study. Lancet (London, England) 2020; 395: 1054–62.

6 Vijayvargiya P, Esquer Garrigos Z, Castillo Almeida NE, Gurram PR, Stevens RW, Razonable RR. Treatment Considerations for COVID-19: A Critical Review of the Evidence (or Lack Thereof). Mayo Clin Proc 2020; 95: 1454–66.

7 Bartoszko JJ, Siemieniuk RAC, Kum E, et al. Prophylaxis against covid-19: living systematic review and network meta-analysis. BMJ 2021; 373: n949.

8 Lee AC, Jeong Y, Lee S, et al. Nasopharyngeal Type-I Interferon for Immediately Available Prophylaxis Against Emerging Respiratory Viral Infections. Front Immunol 2021; 12: 660298.

9 Rodrigues CMC, Plotkin SA. Impact of Vaccines; Health, Economic and Social Perspectives. Front Microbiol 2020; 11: 1526.

10 Kyriakidis NC, López-Cortés A, González EV, Grimaldos AB, Prado EO. SARS-CoV-2 vaccines strategies: a comprehensive review of phase 3 candidates. NPJ vaccines 2021; 6: 28.

11 Gao Q, Bao L, Mao H, et al. Development of an inactivated vaccine candidate for SARS-CoV-2. Science 2020; 369: 77–81.

12 Zhang Y, Zeng G, Pan H, et al. Safety, tolerability, and immunogenicity of an inactivated SARS-CoV-2 vaccine in healthy adults aged 18-59 years: a randomised, double-blind, placebo-controlled, phase 1/2 clinical trial. Lancet Infect Dis 2021; 21: 181–92.

13 Wu Z, Hu Y, Xu M, et al. Safety, tolerability, and immunogenicity of an inactivated SARS-CoV-2 vaccine (CoronaVac) in healthy adults aged 60 years and older: a randomised, double-blind, placebo-controlled, phase 1/2 clinical trial. Lancet Infect Dis 2021; 21: 803–12.

14 Staff R. Sinovac’s coronavirus vaccine candidate approved for emergency use in China –source. 2020. https://www.reuters.com/article/us-health-coronavirus-china-vaccines-idUSKBN25O0Z3 (accessed Jan 31, 2022).

15 Valdespino Gómez JL, García García MDL. Declaración de Helsinki. Gac. Med. Mex. 2001; 137: 391.

16 Bueno SM, Abarca K, González PA, et al. Safety and Immunogenicity of an Inactivated SARS-CoV-2 Vaccine in a Subgroup of Healthy Adults in Chile. Clin Infect Dis 2021; : 2021.03.31.21254494.

17 VACcines SP for E. D2.3 Priority List of Adverse Events of Special Interest: COVID-19. 2020 https://brightoncollaboration.us/wp-content/uploads/2020/06/SPEAC_D2.3_V2.0_COVID-19_20200525_public.pdf (accessed Jan 31, 2022).

18 U. S. Department of Health and Human Services, Services USD of H and H, Administration F and D, Research C for BE and. Toxicity Grading Scale for Healthy Adult and Adolescent Volunteers Enrolled in Preventive Vaccine Clinical Trials. Guid Ind 2007. http://www.fda.gov/cber/guidelines.htm. (accessed Aug 14, 2019).

19 National Cancer Institute. Common Terminology Criteria for Adverse Events (CTCAE) version 5.0. NIH Publ 2017.

20 WHO-UM. The use of the WHO-UMC system for standardized case causality assessment. Uppsala Uppsala Monit. Cent. 2014.

21 WHO. Public Health Surveillance for COVID-19. Interim Guid 2020; : 253–78.

22 Polack FP, Thomas SJ, Kitchin N, et al. Safety and Efficacy of the BNT162b2 mRNA Covid-19 Vaccine. N Engl J Med 2020; 383: 2603–15.

23 Voysey M, Clemens SAC, Madhi SA, et al. Safety and efficacy of the ChAdOx1 nCoV-19 vaccine (AZD1222) against SARS-CoV-2: an interim analysis of four randomised controlled trials in Brazil, South Africa, and the UK. Lancet 2021. DOI:10.1016/S0140-6736(20)32661-1.

24 Barda N, Dagan N, Ben-Shlomo Y, et al. Safety of the BNT162b2 mRNA Covid-19 Vaccine in a Nationwide Setting. N Engl J Med 2021; 385: 1078–90.

25 Palacios R, Batista AP, Albuquerque CSN, et al. Efficacy and Safety of a COVID-19 Inactivated Vaccine in Healthcare Professionals in Brazil: The PROFISCOV Study. SSRN Electron J 2021. DOI:10.2139/ssrn.3822780.

26 Tanriover MD, Doğanay HL, Akova M, et al. Efficacy and safety of an inactivated whole-virion SARS-CoV-2 vaccine (CoronaVac): interim results of a double-blind, randomised, placebo-controlled, phase 3 trial in Turkey. Lancet 2021; 398: 213–22.

27 Zhu F-C, Li Y-H, Guan X-H, et al. Safety, tolerability, and immunogenicity of a recombinant adenovirus type-5 vectored COVID-19 vaccine: a dose-escalation, open-label, non-randomised, first-in-human trial. Lancet (London, England) 2020; 395: 1845–54.

28 Duarte LF, Gálvez NMS, Iturriaga C, et al. Immune Profile and Clinical Outcome of Breakthrough Cases After Vaccination With an Inactivated SARS-CoV-2 Vaccine. Front Immunol 2021; 12: 742914.

29 Jara A, Undurraga EA, González C, et al. Effectiveness of an Inactivated SARS-CoV-2 Vaccine in Chile. N Engl J Med 2021; 385: 875–84.

30 Ho FK, Petermann-Rocha F, Gray SR, et al. Is older age associated with COVID-19 mortality in the absence of other risk factors? General population cohort study of 470,034 participants. PLoS One 2020; 15: e0241824.

