## Supplementary Tables for "Safety and efficacy of two immunization schedules with an inactivated SARS-CoV-2 vaccine in adults. A randomized non-inferiority clinical trial"

|  |  | First dose | |  | Second dose | |
| --- | --- | --- | --- | --- | --- | --- |
|  |  | (n=2302) | |  | (n=2212) | |
|  |  | Schedule 0-14 | Schedule 0-28 |  | Schedule 0-14 | Schedule 0-28 |
|  |  | (n=1090) | (n=1212) |  | (n=1038) | (n=1174) |
| **Local adverse events, n (%)** | |  |  |  |  |  |
|  | Pain | 22 (2.02) | 17 (1.4) |  | 22 (2.12) | 11 (0.94) |
|  | Induration | 0 (0.00) | 1 (0.08) |  | 0 (0.00) | 2 (0.17) |
|  | Pruritus | 3 (0.28) | 0 (0.00) |  | 5 (0.48) | 1 (0.09) |
|  | Redness | 3 (0.28) | 1 (0.08) |  | 2 (0.19) | 1 (0.09) |
|  | Swelling | 0 (0.00) | 0 (0.00) |  | 0 (0.00) | 1 (0.09) |
| **Systemic adverse events, n (%)** | |  |  |  |  |  |
|  | Headache | 6 (0.55) | 6 (0.5) |  | 4 (0.39) | 3 (0.26) |
|  | Fatigue | 1 (0.09) | 1 (0.08) |  | 2 (0.19) | 2 (0.17) |
|  | Muscle pain | 0 (0.00) | 1 (0.08) |  | 0 (0.00) | 1 (0.09) |
|  | Diarrhoea | 1 (0.09) | 1 (0.08) |  | 0 (0.00) | 1 (0.09) |
|  | Nausea | 0 (0.00) | 1 (0.08) |  | 1 (0.10) | 2 (0.17) |
|  | Anorexia | 0 (0.00) | 0 (0.00) |  | 0 (0.00) | 1 (0.09) |
|  | Arthralgia | 0 (0.00) | 0 (0.00) |  | 0 (0.00) | 0 (0.00) |
|  | Pruritus | 2 (0.18) | 0 (0.00) |  | 1 (0.10) | 0 (0.00) |
|  | Exanthema | 0 (0.00) | 0 (0.00) |  | 0 (0.00) | 0 (0.00) |
|  | Allergic reaction | 0 (0.00) | 0 (0.00) |  | 0 (0.00) | 0 (0.00) |
|  | Vomiting | 0 (0.00) | 0 (0.00) |  | 0 (0.00) | 0 (0.00) |
|  | Fever (>37.8ºC) | 0 (0.00) | 0 (0.00) |  | 0 (0.00) | 0 (0.00) |

Supplementary Table 1. Frequency of adverse events within first 60 min post vaccine administration, by dose and schedule.

Supplementary Table 2. Frequency of non-immediate local and systemic adverse events by dose and schedule.

|  |  |  | First dose | |  | Second dose | | |
| --- | --- | --- | --- | --- | --- | --- | --- | --- |
|  |  |  | (n=2302) | |  | (n=2212) | | |
|  |  |  | Schedule 0-14 | Schedule 0-28 |  | Schedule 0-14 | Schedule 0-28 | *Non-inferiority* |
|  |  |  | (n=1090) | (n=1212) |  | (n=1038) | (n=1174) | *p value (a)* |
| **Local adverse events,** n (%) | |  |  |  |  |  |  |  |
|  | Pain |  | 347 (31.8) | 351 (29) |  | 335 (32.3) | 316 (26.9) | <0.001 |
|  | Induration |  | 22 (2.0) | 36 (3.0) |  | 52 (5.0) | 27 (2.3) | <0.001 |
|  | Pruritus |  | 28 (2.6) | 21 (1.7) |  | 32 (3.1) | 13 (1.1) | <0.001 |
|  | Redness |  | 27 (2.5) | 14 (1.2) |  | 24 (2.3) | 11 (0.9) | <0.001 |
|  | Swelling |  | 17 (1.6) | 19 (1.6) |  | 31 (3.0) | 26 (2.2) | <0.001 |
| **Systemic adverse events,** n (%) | | |  |  |  |  |  |  |
|  | Headache |  | 282 (25.9) | 311 (25.7) |  | 224 (21.6) | 236 (20.1) | <0.001 |
|  | Fatigue |  | 173 (15.9) | 209 (17.2) |  | 122 (11.8) | 149 (12.7) | <0.001 |
|  | Muscle pain |  | 150 (13.8) | 172 (14.2) |  | 113 (10.9) | 135 (11.5) | <0.001 |
|  | Diarrhoea |  | 106 (9.7) | 95 (7.8) |  | 64 (6.2) | 73 (6.2) | <0.001 |
|  | Nausea |  | 62 (5.7) | 69 (5.7) |  | 39 (3.8) | 48 (4.1) | <0.001 |
|  | Anorexia |  | 51 (4.7) | 47 (3.9) |  | 19 (1.8) | 24 (2.0) | <0.001 |
|  | Arthralgia |  | 38 (3.5) | 51 (4.2) |  | 22 (2.1) | 44 (3.7) | <0.001 |
|  | Pruritus |  | 23 (2.1) | 12 (1.0) |  | 18 (1.7) | 8 (0.7) | <0.001 |
|  | Exanthema |  | 19 (1.7) | 11 (0.9) |  | 8 (0.8) | 8 (0.7) | <0.001 |
|  | Allergic reaction |  | 15 (1.4) | 7 (0.6) |  | 8 (0.8) | 9 (0.8) | <0.001 |
|  | Vomiting |  | 5 (0.5) | 5 (0.4) |  | 7 (0.7) | 10 (0.9) | <0.001 |
|  | Fever (>37.8ºC) |  | 2 (0.2) | 4 (0.3) |  | 3 (0.3) | 4 (0.3) | <0.001 |
| (a) Non-inferiority test, with a margin of 15%, for comparing schedule 0-14 against 0-28. | | | | | | | | |

Supplementary Table 3. Number of simultaneous not-immediate adverse events post vaccine administration by dose and schedule.

|  |  | First dose | |  | Second dose | |
| --- | --- | --- | --- | --- | --- | --- |
|  |  | (n=2302) | |  | (n=2212) | |
|  |  | Schedule 0-14 | Schedule 0-28 |  | Schedule 0-14 | Schedule 0-28 |
|  |  | (n=1090) | (n=1212) |  | (n=1038) | (n=1174) |
| **Local adverse events, n (%)** | |  |  |  |  |  |
|  | 0 | 746 (68.4) | 857 (70.7) |  | 700 (67.4) | 857 (73.0) |
|  | 1 | 304 (27.9) | 305 (25.2) |  | 276 (26.6) | 276 (23.5) |
|  | 2 | 23 (2.1) | 37 (3.1) |  | 34 (3.3) | 27 (2.3) |
|  | 3 | 12 (1.1) | 10 (0.8) |  | 17 (1.6) | 10 (0.9) |
|  | ≥4 | 5 (0.5) | 3 (0.2) |  | 11 (1.1) | 4 (0.3) |
| **Systemic adverse events, n (%)** | |  |  |  |  |  |
|  | 0 | 643 (59.0) | 709 (58.5) |  | 683 (65.8) | 808 (68.8) |
|  | 1 | 208 (19.1) | 245 (20.2) |  | 190 (18.3) | 187 (15.9) |
|  | 2 | 113 (10.4) | 136 (11.2) |  | 86 (8.3) | 80 (6.8) |
|  | 3 | 68 (6.2) | 63 (5.2) |  | 54 (5.2) | 44 (3.7) |
|  | ≥4 | 58 (5.3) | 59 (4.9) |  | 25 (2.4) | 55 (4.7) |

| Supplementary Table 4. Duration of adverse events by dose and schedule. | | | | | | | | | | |
| --- | --- | --- | --- | --- | --- | --- | --- | --- | --- | --- |
|  |  |  | First dose | | |  |  | Second dose | | |
|  |  | Schedule 0-14 | | Schedule 0-28 | |  | Schedule 0-14 | | Schedule 0-28 | |
|  |  | n | Days, median  (P10-P90) | n | Days, median  (P10-P90) |  | n | Days, median  (P10-P90) | n | Days, median  (P10-P90) |
| **Local adverse events** | |  |  |  |  |  |  |  |  |  |
|  | Pain | 347 | 2 (1 - 3) | 351 | 1 (1 - 3) |  | 335 | 2 (1 - 4) | 316 | 2 (1 - 4) |
| **Systemic adverse events** | |  |  |  |  |  |  |  |  |  |
|  | Headache | 282 | 2 (1 - 3) | 311 | 1 (1 - 4) |  | 224 | 1 (1 - 4) | 236 | 2 (1 - 5) |
|  | Fatigue | 173 | 2 (1 - 4) | 209 | 1 (1 - 3) |  | 122 | 2 (1 - 4) | 149 | 2 (1 - 4) |
|  | Muscle pain | 150 | 2 (1 - 4) | 172 | 1 (1 - 4) |  | 113 | 2 (1 - 5) | 135 | 2 (1 - 5) |
| P10: 10th quantile; P90: 90th quantile | | | | | | | | | | |

Supplementary Table 5. Frequency of adverse events by schedule, dose and age group.

|  |  |  | First dose | |  | Second dose | |
| --- | --- | --- | --- | --- | --- | --- | --- |
|  |  |  | (n=2302) | |  | (n=2212) | |
|  |  |  | Schedule 0-14 | Schedule 0-28 |  | Schedule 0-14 | Schedule 0-28 |
|  |  |  | (n=1090) | (n=1212) |  | (n=1038) | (n=1174) |
| **Local adverse events** | |  |  |  |  |  |  |
| Pain; n (%) | |  |  |  |  |  |  |
|  | <60 years |  | 303 (37.9) | 305 (37.4) |  | 296 (39.3) | 265 (33.5) |
|  | >=60 years |  | 44 (15.2) | 46 (11.6) |  | 39 (13.7) | 51 (13.3) |
|  | *p value (a)* |  | ***<0.001*** | ***<0.001*** |  | ***<0.001*** | ***<0.001*** |
| Induration; n (%) | |  |  |  |  |  |  |
|  | <60 years |  | 18 (2.3) | 25 (3.1) |  | 45 (6.0) | 22 (2.8) |
|  | >=60 years |  | 4 (1.4) | 11 (2.8) |  | 7 (2.5) | 5 (1.3) |
|  | *p value (a)* |  | *0.366* | *0.783* |  | *0.021* | *0.112* |
| Pruritus; n (%) | |  |  |  |  |  |  |
|  | <60 years |  | 26 (3.3) | 19 (2.3) |  | 27 (3.6) | 12 (1.5) |
|  | >=60 years |  | 2 (0.7) | 2 (0.5) |  | 5 (1.8) | 1 (0.3) |
|  | *p value (a)* |  | *0.018* | *0.023* |  | *0.130* | *0.072* |
| Redness; n (%) | |  |  |  |  |  |  |
|  | <60 years |  | 23 (2.9) | 12 (1.5) |  | 21 (2.8) | 9 (1.1) |
|  | >=60 years |  | 4 (1.4) | 2 (0.5) |  | 3 (1.1) | 2 (0.5) |
|  | *p value (a)* |  | *0.160* | *0.140* |  | *0.098* | *0.519* |
| Swelling; n (%) | |  |  |  |  |  |  |
|  | <60 years |  | 14 (1.8) | 12 (1.5) |  | 24 (3.2) | 17 (2.2) |
|  | >=60 years |  | 3 (1.0) | 7 (1.8) |  | 7 (2.5) | 9 (2.3) |
|  | *p value (a)* |  | *0.400* | *0.696* |  | *0.544* | *0.834* |
| **Systemic adverse reactions** | |  |  |  |  |  |  |
| Headache; n (%) | |  |  |  |  |  |  |
|  | <60 years |  | 236 (29.5) | 243 (29.8) |  | 180 (23.9) | 186 (23.5) |
|  | >=60 years |  | 46 (15.9) | 68 (17.2) |  | 44 (15.5) | 50 (13.0) |
|  | *p value (a)* |  | ***<0.001*** | ***<0.001*** |  | *0.003* | *<0.001* |
| Fatigue; n (%) | |  |  |  |  |  |  |
|  | <60 years |  | 145 (18.1) | 163 (20.0) |  | 92 (12.2) | 111 (14.1) |
|  | >=60 years |  | 28 (9.7) | 46 (11.6) |  | 30 (10.6) | 38 (9.9) |
|  | *p value (a)* |  | ***0.001*** | ***<0.001*** |  | *0.465* | *0.045* |
| Muscle pain; n (%) | |  |  |  |  |  |  |
|  | <60 years |  | 125 (15.6) | 138 (16.9) |  | 78 (10.3) | 103 (13.0) |
|  | >=60 years |  | 25 (8.6) | 34 (8.6) |  | 35 (12.3) | 32 (8.3) |
|  | *p value (a)* |  | *0.003* | ***<0.001*** |  | *0.361* | *0.018* |
| Diarrhoea; n (%) | |  |  |  |  |  |  |
|  | <60 years |  | 87 (10.9) | 77 (9.4) |  | 56 (7.4) | 61 (7.7) |
|  | >=60 years |  | 19 (6.6) | 18 (4.5) |  | 8 (2.8) | 12 (3.1) |
|  | *p value (a)* |  | *0.033* | *0.003* |  | ***0.006*** | *0.002* |
| Nausea; n (%) | |  |  |  |  |  |  |
|  | <60 years |  | 54 (6.8) | 55 (6.7) |  | 33 (4.4) | 34 (4.3) |
|  | >=60 years |  | 8 (2.8) | 14 (3.5) |  | 6 (2.1) | 14 (3.6) |
|  | *p value (a)* |  | *0.012* | *0.024* |  | *0.087* | *0.593* |
| Anorexia; n (%) | |  |  |  |  |  |  |
|  | <60 years |  | 47 (5.9) | 43 (5.3) |  | 16 (2.1) | 17 (2.2) |
|  | >=60 years |  | 4 (1.4) | 4 (1.0) |  | 3 (1.1) | 7 (1.8) |
|  | *p value (a)* |  | ***0.002*** | *<0.001* |  | *0.254* | *0.709* |
| Athralgia; n (%) | |  |  |  |  |  |  |
|  | <60 years |  | 30 (3.8) | 35 (4.3) |  | 14 (1.9) | 28 (3.5) |
|  | >=60 years |  | 8 (2.8) | 16 (4.0) |  | 8 (2.8) | 16 (4.2) |
|  | *p value (a)* |  | *0.43* | *0.840* |  | *0.338* | *0.598* |
| Pruritus; n (%) | |  |  |  |  |  |  |
|  | <60 years |  | 18 (2.3) | 11 (1.3) |  | 15 (2.0) | 5 (0.6) |
|  | >=60 years |  | 5 (1.7) | 1 (0.3) |  | 3 (1.1) | 3 (0.8) |
|  | *p value (a)* |  | *0.593* | *0.117* |  | *0.427* | *0.721* |
| Exanthema; n (%) | |  |  |  |  |  |  |
|  | <60 years |  | 16 (2.0) | 7 (0.9) |  | 6 (0.8) | 3 (0.4) |
|  | >=60 years |  | 3 (1.0) | 4 (1.0) |  | 2 (0.7) | 5 (1.3) |
|  | *p value (a)* |  | *0.282* | *0.756* |  | *0.999* | *0.123* |
| Allergic reaction; n (%) | |  |  |  |  |  |  |
|  | <60 years |  | 13 (1.6) | 5 (0.6) |  | 6 (0.8) | 6 (0.8) |
|  | >=60 years |  | 2 (0.7) | 2 (0.5) |  | 2 (0.7) | 3 (0.8) |
|  | *p value (a)* |  | *0.378* | *0.999* |  | *0.999* | *0.999* |
| Vomiting; n (%) | |  |  |  |  |  |  |
|  | <60 years |  | 4 (0.5) | 4 (0.5) |  | 6 (0.8) | 6 (0.8) |
|  | >=60 years |  | 1 (0.3) | 1 (0.3) |  | 1 (0.4) | 4 (1.0) |
|  | *p value (a)* |  | *0.738* | *0.999* |  | *0.681* | *0.737* |
| Fever (>37.8ºC); n (%) | |  |  |  |  |  |  |
|  | <60 years |  | 2 (0.3) | 3 (0.4) |  | 3 (0.4) | 4 (0.5) |
|  | >=60 years |  | 0 (0.0) | 1 (0.3) |  | 0 (0.0) | 0 (0.0) |
|  | *p value (a)* |  | *0.999* | *0.999* |  | *0.566* | *0.310* |
| Sample sizes for first dose. Schedule 0-14 <60 years n=800; schedule 0-14 >=60 years n=290; schedule 0-28 <60 years n=816; schedule 0-28 >=60 years n=396. Sample sizes for second dose. Schedule 0-14 <60 years n=754; schedule 0-14 >=60 years n=284; schedule 0-28 <60 years n=790; schedule 0-28 >=60 years n=384. | | | | | | | |
| (a) For each scheme subgroup, AE differences by age group were compared using two-sided tests. | | | | | | | |

Supplementary Table 6. Incidence of COVID-19 positive patients by immunization schedule in different subpopulations.

|  | | Schedule 0-14 | | Schedule 0-28 | |  |
| --- | --- | --- | --- | --- | --- | --- |
|  |  | N  Total | % COVID19 | N  Total | %  COVID19 | Non-inferiority  p value* |
| Age (years) | |  |  |  |  |  |
|  | 18-59 | 760 | 3.9 | 793 | 2.4 | <0.001 |
|  | 60-98 | 285 | 1.8 | 387 | 1.3 | <0.001 |
| Sex | |  |  |  |  |  |
|  | Female | 751 | 3.2 | 588 | 2.2 | <0.001 |
|  | Male | 465 | 3.4 | 583 | 1.9 | <0.001 |
| Health setting workers | |  |  |  |  |  |
|  | Yes | 421 | 2.4 | 290 | 1.7 | <0.001 |
|  | No | 615 | 4.5 | 881 | 3.1 | <0.001 |
| Comorbidities | |  |  |  |  |  |
|  | >=1 | 465 | 3.0 | 531 | 2.3 | <0.001 |
|  | None | 571 | 3.5 | 639 | 1.9 | <0.001 |
| Obesity | |  |  |  |  |  |
|  | Yes | 194 | 2.6 | 261 | 3.4 | <0.001 |
|  | No | 841 | 3.4 | 909 | 1.7 | <0.001 |
| Incidence of COVID-19 14 days post second dose or after. | | | | | | |
| Data presented as sample size for each schedule segmented by levels of sociodemographic characteristic and percentage of COVID-19 cases in each subgroup. *Non-inferiority test, with a margin of 15%, for comparing schedule 0-14 against 0-28. | | | | | | |
| (a) Two asymptomatic cases, three COVID-19 cases before 14 days post 2nd dose, and those inoculated out of vaccination window were omitted from this analysis. | | | | | | |
